# Data-driven subtypes of type 2 diabetes mellitus and risk of dementia, stroke, and brain structural changes in the UK Biobank

**DOI:** 10.64898/2026.03.30.26349725

**Authors:** Si Han, Yingying Zhou, Miriam Sturkenboom, Geert Jan Biessels, Fariba Ahmadizar

## Abstract

**Aims:** Type 2 diabetes mellitus (T2DM) increases risks of stroke and dementia, yet these risks vary across individuals. We hypothesized that clinically derived diabetes subtypes contribute to this heterogeneity. We aimed to identify data-driven subtypes using routine clinical features and examine their associations with dementia, stroke, mortality, and brain structure.

**Methods:** K-means clustering was applied to 14,353 UK Biobank participants with prevalent T2DM using age at diagnosis, body mass index, glycated hemoglobin, insulin resistance (triglyceride/HDL ratio), systolic blood pressure, and C-reactive protein. Cox models assessed associations with incident dementia (all-cause, Alzheimer’s disease [AD], vascular dementia [VaD]), stroke (all-cause, ischemic [IS], intracerebral hemorrhage [ICH]), and mortality. Brain MRI outcomes were analyzed in 779 participants using inverse probability-weighted linear regression.

**Results:** Three subtypes were identified: severe obesity-related inflammatory diabetes (SOID), mild metabolic diabetes (MMD, reference), and mild age-related hypertension-predominant diabetes (MARD-H). Compared with MMD, SOID showed higher risks of dementia (HR 1.24), VaD (HR 1.42), stroke (HR 1.38), IS (HR 1.48), all-cause mortality (HR 1.59), and cardiovascular death (HR 1.88). MRI showed lower gray matter volume and greater white matter hyperintensity burden in SOID.

**Conclusions:** Data-driven subtyping revealed heterogeneity in neurological risk in T2DM, with the obesity-inflammation subtype showing elevated vascular and neuroimaging risk.

**Research in Context:** *What is already known about this subject?:*

- Data-driven subtypes of type 2 diabetes mellitus using clinical or genetic features have revealed substantial heterogeneity.
- Type 2 diabetes mellitus is associated with increased risks of stroke and dementia, particularly vascular dementia, yet it remains unclear whether these neurological outcomes vary across clinically derived type 2 diabetes mellitus subtypes.

*What is the key question?:*

- Do type 2 diabetes mellitus subtypes data-derived from routinely measured clinical variables show differential associations with stroke and dementia subtypes, and brain structural measures?

*What are the new findings?:*

- Three type 2 diabetes mellitus subtypes were identified in a large population-based cohort using routinely collected data.
- Severe obesity-related inflammatory diabetes subtype showed a higher risk of vascular dementia, ischemic stroke, mortality, and greater white matter hyperintensity burden, whereas associations with Alzheimer’s disease were not observed.

*How might this impact clinical practice in the foreseeable future?:*

- Clinical subtyping of type 2 diabetes mellitus may help characterize heterogeneity in neurological risk and identify patient groups with disproportionate vulnerability to vascular brain injury, although further validation is required before clinical implementation.

## Introduction

Type 2 diabetes mellitus (T2DM) affects more than 530 million adults worldwide and is a major contributor to morbidity and mortality(1). In addition to its well-established cardiovascular and renal complications, T2DM is associated with increased risks of dementia, particularly vascular dementia, and stroke, which together represent a substantial and growing public health burden (2–5). However, these neurological risks are not uniform across individuals with T2DM, suggesting heterogeneity in the underlying pathways linking metabolic dysfunction to cerebrovascular and neurodegenerative outcomes.

Recent data-driven clustering studies have identified distinct T2DM subtypes based on routinely measured clinical and metabolic features, such as age at diagnosis, adiposity, glycemic control, and insulin-resistant (6–8). These subtypes differ in their risks of micro-and macrovascular complications (9, 10), supporting the concept that T2DM is a heterogeneous condition with distinct cardiometabolic trajectories. Whether such clinically derived T2DM subtypes also differ in their risks of specific neurological outcomes, including dementia subtypes, stroke subtypes, and structural brain changes, remain insufficiently understood.

In this study, we applied an unsupervised machine-learning clustering approach using routinely collected clinical variables to drive data-driven T2DM subtypes in a large population-based cohort. We examined their associations with incident dementia (including Alzheimer’s disease [AD] and vascular dementia [VaD]), stroke subtypes (all-cause, ischemic [IS], and hemorrhagic [ICH]), cause-specific mortality, and brain MRI measures. By integrating clinical outcomes with brain structural markers, this study aims to clarify the heterogeneity of neurological risk among individuals with T2DM and to identify subgroups with disproportionate vulnerability to vascular brain injury.

## Methods

### 2.1 Study Population

The UK Biobank (UKB) is a large, population-based prospective cohort, consisting of >500,000 participants aged 40-69 years at baseline recruitment in 2006-2010. Participants completed a touchscreen questionnaire and a verbal interview, took physical measurements, and provided biological samples (blood, urine, and saliva) in 1 of 22 assessment centers throughout England, Scotland, and Wales. The UKB received ethics approval from the Northwest Multicenter Research Ethics Committee (reference no. 16/NW/0274). All participants provided written informed consent for the study.

Participants with diabetes at baseline were identified using multiple data sources, consistent with previous studies (11, 12). Self-reported medical history was collected via touchscreen questionnaire or verbal interview during the baseline assessment. Prevalent diabetes was identified using a previously defined algorithm (12). Hospital admissions records were obtained through linkage to Health Episode Statistics (England and Wales) and Scottish Morbidity Records (Scotland). T2DM identified from hospital records was classified using ICD-9 codes 250.00, 250.10, 250.20, and 250.90 or ICD-10 code E11. Participants with self-reported or hospital diagnoses were identified as diagnosed cases. Undiagnosed T2DM patients were defined according to the American Diabetes Association criteria as random glucose ≥11.1 mmol/L or glycated hemoglobin (HbA1c) ≥48 mmol/mol (6.5%). In total, 14,353 participants with T2DM were included in the analysis (*Supplementary Figure S1*).

### 2.2 Outcomes

Primary outcomes were incident dementia (all-cause, AD, and VaD) and stroke (all-cause, IS, and ICH), in the period after T2DM diagnosis. Outcomes were defined using validated UKB algorithms integrating hospital inpatient records, primary care, and mortality records (13) (*Supplementary Table S2-S3)*. Censoring dates were defined as the last day of the most recent month for which data were estimated to be largely complete (>90% of the average number of records from the preceding three months).

Secondary outcomes included MRI-derived brain structural measures: total brain, gray matter, white matter, hippocampal, and white matter hyperintensity (WMH) volumes, normalized for intracranial volume where available. Brain MRI data were obtained from the UKB imaging study initiated in 2015. Details of imaging acquisition, processing, and quality control are provided in *Supplementary Text S1*. MRI data were available for ∼5% of participants and analyzed as secondary outcomes in this subset.

Mortality data were obtained from national death registries in England, Scotland, and Wales to define all-cause and cause-specific mortality (cardiovascular and non-cardiovascular). Follow-up time was calculated from the date of T2DM diagnosis to the first occurrence of the outcome, death, loss to follow-up, or the censoring date, whichever came first. Primary follow-up time was defined from the baseline assessment, when cluster-defining variables were measured. For participants diagnosed before baseline, delayed entry was applied to account for time between diagnosis and study entry.

### 2.3 Covariates

Covariates were established risk factors for cognitive, vascular, and metabolic health. A simplified directed acyclic graph (DAG) illustrating the hypothesized relationships between T2DM–related metabolic profiles and neurological outcomes informed covariate selection (*Supplementary Figure S2)*.

Demographic covariates included age at baseline, sex, ethnicity, educational attainment, and the Townsend deprivation index. Lifestyle-related factors comprised smoking status, alcohol consumption, and an overall diet quality score reflecting habitual dietary intake during the year preceding the baseline assessment. The estimated glomerular filtration rate (eGFR) was calculated using the CKD-EPI 2021 equation. Baseline use of lipid-lowering, blood pressure-lowering, and glucose-lowering medications was considered a downstream correlate of diabetes severity and was included in additional adjustment models to assess the robustness of associations.

### 2.4 Cluster Analysis

Six routinely measured clinical variables reflecting metabolic status, cardiovascular risk, and systemic inflammation were used to derive T2DM subtypes: age at diagnosis, body mass index (BMI), glycated hemoglobin (HbA1c), insulin resistance (IR) estimated using the triglyceride-to–HDL cholesterol ratio, systolic blood pressure (SBP), and C-reactive protein (CRP). Prior to clustering, missing values in the cluster-defining variables were handled using random forest imputation. Extreme outliers exceeding ±5 standard deviations (SD) were excluded. Right-skewed variables were log-transformed to reduce skewness. All six variables were then standardized to z-scores to ensure equal weighting in the clustering process. Unsupervised k-means clustering was performed using 50-100 random initializations to minimize convergence to local minima. The optimal number of clusters (k = 2-8) was determined via the elbow method based on within-cluster sum of squares (WSS). To assess structural robustness, hierarchical clustering using Ward’s D^2^ linkage was applied in parallel. Cluster stability was further evaluated using nonparametric bootstrap resampling with 200 replicates. All variables were obtained at or near the baseline assessment visit, defined as within one year of T2DM ascertainment. When multiple measurements were available within this window, the value closest to baseline was used. Laboratory assays and measurement procedures followed standard UKB protocols, with detailed assay information, device specifications, and unit conversions provided in *Supplementary Text S1*.

### 2.5 Outcome analysis

Associations between T2DM subtypes and incident dementia or stroke were assessed using Cox proportional hazards models. Hazard ratios (HRs) and 95% confidence intervals (CIs) were estimated using the metabolically mild cluster as the reference group, selected a priori because it represented the most common and clinically least severe subtype.

To complement relative estimates, 10-year cumulative incidence and restricted mean survival time (RMST) were calculated for each cluster. The cumulative incidence function (CIF) estimated the absolute 10-year probability of the outcome, while RMST represented the mean event-free survival time over 10 years.

Covariates were added sequentially in four nested models: Model 1 (unadjusted), Model 2 (adjusted for age at baseline, sex, ethnicity), Model 3 (further adjusted for education, deprivation index, smoking status, alcohol consumption, and diet quality); and Model 4 (additionally adjusted for kidney function and medication use). Model 3 was considered the primary inferential model, while Model 4 was interpreted cautiously due to potential overadjustment for downstream correlates of diabetes severity.

All-cause and cause-specific mortality were analyzed using Cox proportional hazards models, with the proportional hazards assumption assessed using Schoenfeld residuals. To account for non-random MRI participation (n = 779, 5.43%), each individual’s probability of undergoing brain MRI was estimated using a logistic selection model including age, sex, ethnicity, education, deprivation index, and lifestyle factors. Weighted linear regression models were fitted to assess associations between clusters and standardized MRI outcomes, including total brain, white matter, gray matter, hippocampal volume, and log-transformed WMH volume. Results are reported as standardized β coefficients with 95% CIs.

To control for multiple comparisons across all the outcomes, we applied Benjamini-Hochberg false discovery rate (FDR) correction. All statistical analyses were performed using R version 4.5.1 (R Foundation for Statistical Computing, Vienna, Austria) using the survival, fpc, and mice packages.

### 2.6 Sensitivity analyses

Several sensitivity analyses were conducted. First, clustering and outcome analysis were repeated in a complete-case dataset to assess robustness to missing data. Second, to reduce potential reverse causation arising from the long development phase of dementia, we repeated all dementia analyses after excluding participants who developed dementia during the first 5 years of follow-up. Third, we restricted the sample to participants with diabetes duration ≤1 year at baseline to reduce potential survivor bias related to long-standing prevalent disease. The same fully specified models were applied as in the primary analyses. Finally, given the high mortality risk in this population, dementia and stroke analyses were additionally evaluated using cause-specific hazard models treating death as a competing event. Cumulative incidence functions were estimated accounting for death as a competing risk. Fine-Gray subdistribution hazard models were conducted as sensitivity analyses to assess robustness of findings. Multicollinearity was assessed using variance inflation factors (VIFs); diagnostics indicated no covariate exceeded accepted thresholds.

## Results

### 3.1 Cluster characteristics

Among 14,353 participants with T2DM, 5,395 were female, and the mean age was 59.8 years. Baseline demographic and clinical characteristics differed significantly across clusters (all p < 0.001; *Table 1*). Internal validation metrics supported the three-cluster solution. The elbow methods identified k = 3 as optimal, and bootstrap resampling yielded mean Jaccard coefficients ranging from 0.88 to 0.92. Detailed clustering diagnostics are provided in *Supplementary Figures S3-S5*.

**Table 1.**
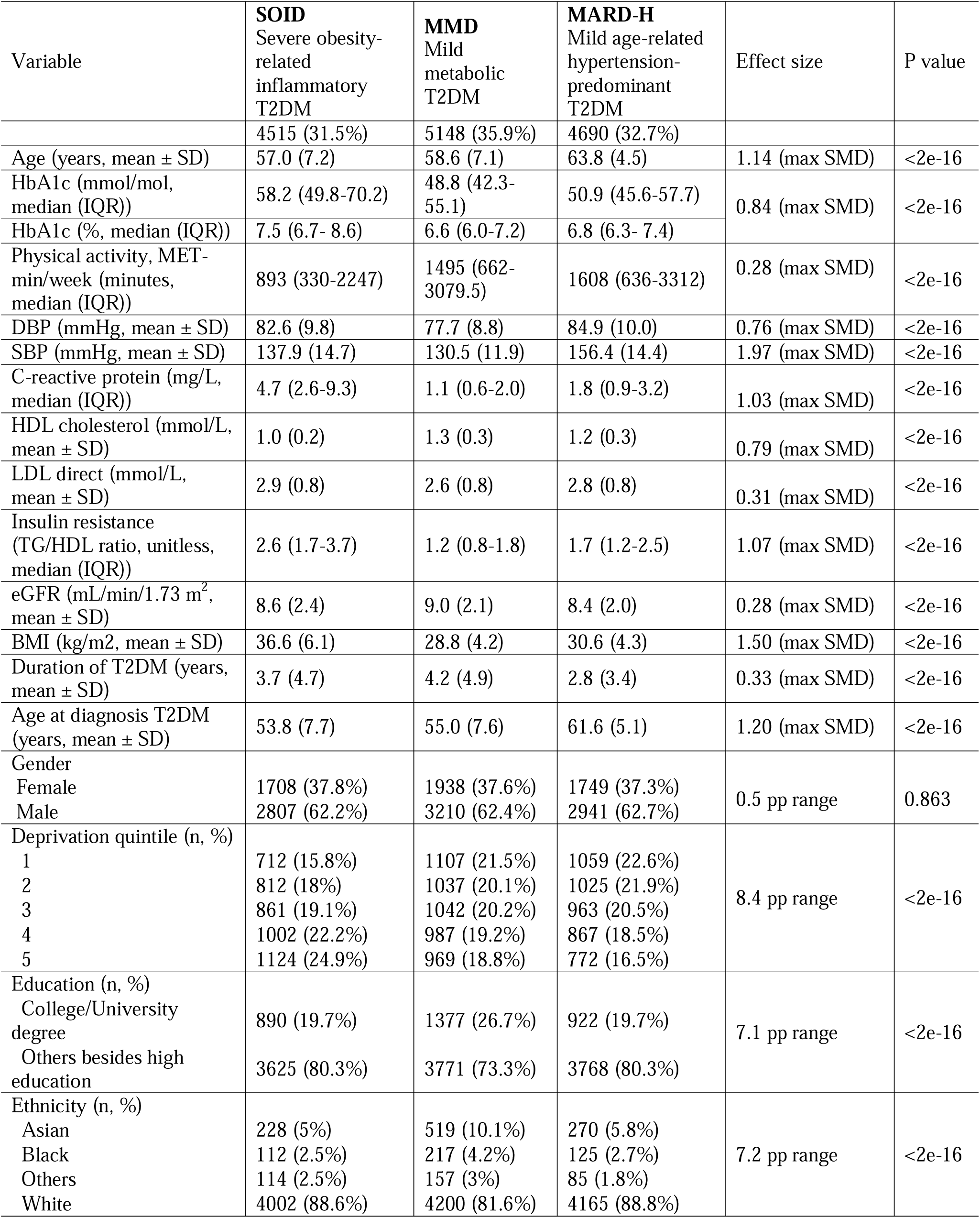
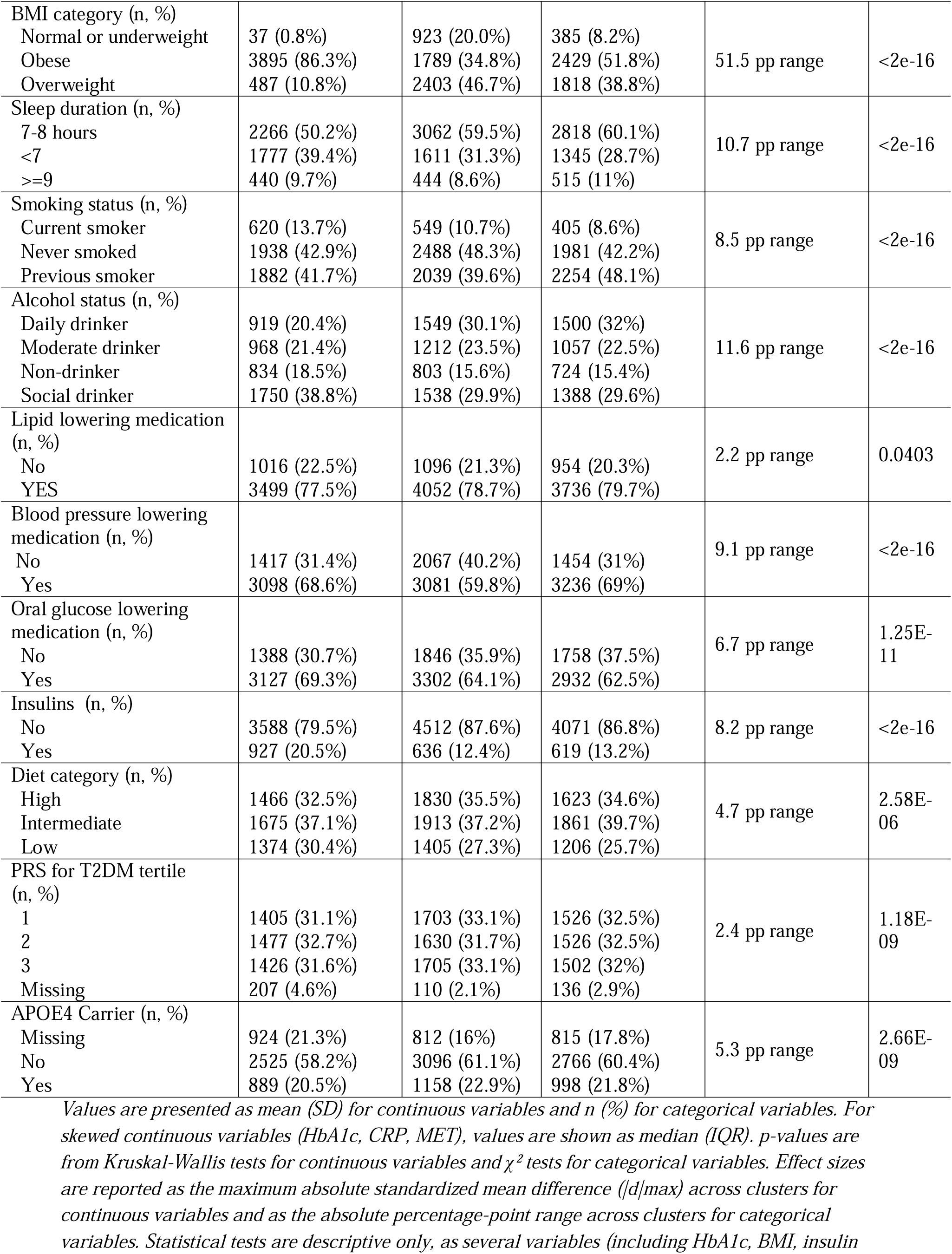
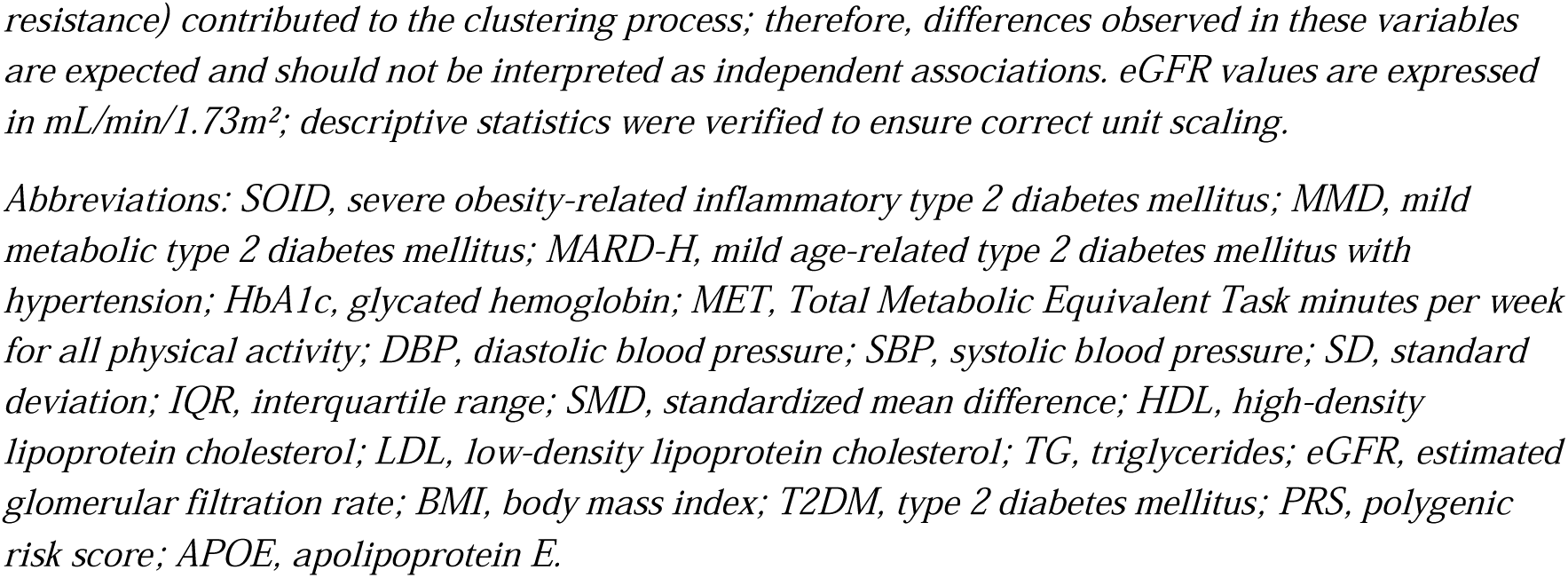
Baseline clinical and metabolic characteristics of the three data-driven type 2 diabetes mellitus subtypes.

| Variable | **SOID**<br>Severe obesity-related<br>inflammatory<br>T2DM | **MMD**<br>Mild<br>metabolic<br>T2DM | **MARD-H**<br>Mild age-related<br>hypertension-predominant<br>T2DM | Effect size | P value |
| --- | --- | --- | --- | --- | --- |
|  | 4515 (31.5%) | 5148 (35.9%) | 4690 (32.7%) |  |  |
| Age (years, mean ± SD) | 57.0 (7.2) | 58.6 (7.1) | 63.8 (4.5) | 1.14 (max SMD) | <2e-16 |
| HbA1c (mmol/mol,<br>median (IQR)) | 58.2 (49.8-70.2) | 48.8 (42.3-55.1) | 50.9 (45.6-57.7) | 0.84 (max SMD) | <2e-16 |
| HbA1c (% , median (IQR)) | 7.5 (6.7- 8.6) | 6.6 (6.0-7.2) | 6.8 (6.3- 7.4) |  |  |
| Physical activity, MET-min/week (minutes,<br>median (IQR)) | 893 (330-2247) | 1495 (662-3079.5) | 1608 (636-3312) | 0.28 (max SMD) | <2e-16 |
| DBP (mmHg, mean ± SD) | 82.6 (9.8) | 77.7 (8.8) | 84.9 (10.0) | 0.76 (max SMD) | <2e-16 |
| SBP (mmHg, mean ± SD) | 137.9 (14.7) | 130.5 (11.9) | 156.4 (14.4) | 1.97 (max SMD) | <2e-16 |
| C-reactive protein (mg/L,<br>median (IQR)) | 4.7 (2.6-9.3) | 1.1 (0.6-2.0) | 1.8 (0.9-3.2) | 1.03 (max SMD) | <2e-16 |
| HDL cholesterol (mmol/L,<br>mean ± SD) | 1.0 (0.2) | 1.3 (0.3) | 1.2 (0.3) | 0.79 (max SMD) | <2e-16 |
| LDL direct (mmol/L,<br>mean ± SD) | 2.9 (0.8) | 2.6 (0.8) | 2.8 (0.8) | 0.31 (max SMD) | <2e-16 |
| Insulin resistance<br>(TG/HDL ratio, unitless,<br>median (IQR)) | 2.6 (1.7-3.7) | 1.2 (0.8-1.8) | 1.7 (1.2-2.5) | 1.07 (max SMD) | <2e-16 |
| eGFR (mL/min/1.73 m<sup>2</sup>,<br>mean ± SD) | 8.6 (2.4) | 9.0 (2.1) | 8.4 (2.0) | 0.28 (max SMD) | <2e-16 |
| BMI (kg/m<sup>2</sup>, mean ± SD) | 36.6 (6.1) | 28.8 (4.2) | 30.6 (4.3) | 1.50 (max SMD) | <2e-16 |
| Duration of T2DM (years,<br>mean ± SD) | 3.7 (4.7) | 4.2 (4.9) | 2.8 (3.4) | 0.33 (max SMD) | <2e-16 |
| Age at diagnosis T2DM<br>(years, mean ± SD) | 53.8 (7.7) | 55.0 (7.6) | 61.6 (5.1) | 1.20 (max SMD) | <2e-16 |
| Gender |  |  |  |  |  |
| Female | 1708 (37.8%) | 1938 (37.6%) | 1749 (37.3%) | 0.5 pp range | 0.863 |
| Male | 2807 (62.2%) | 3210 (62.4%) | 2941 (62.7%) |  |  |
| Deprivation quintile (n, %) |  |  |  |  |  |
| 1 | 712 (15.8%) | 1107 (21.5%) | 1059 (22.6%) | 8.4 pp range | <2e-16 |
| 2 | 812 (18%) | 1037 (20.1%) | 1025 (21.9%) |  |  |
| 3 | 861 (19.1%) | 1042 (20.2%) | 963 (20.5%) |  |  |
| 4 | 1002 (22.2%) | 987 (19.2%) | 867 (18.5%) |  |  |
| 5 | 1124 (24.9%) | 969 (18.8%) | 772 (16.5%) |  |  |
| Education (n, %) |  |  |  |  |  |
| College/University<br>degree | 890 (19.7%) | 1377 (26.7%) | 922 (19.7%) | 7.1 pp range | <2e-16 |
| Others besides high<br>education | 3625 (80.3%) | 3771 (73.3%) | 3768 (80.3%) |  |  |
| Ethnicity (n, %) |  |  |  |  |  |
| Asian | 228 (5%) | 519 (10.1%) | 270 (5.8%) | 7.2 pp range | <2e-16 |
| Black | 112 (2.5%) | 217 (4.2%) | 125 (2.7%) |  |  |
| Others | 114 (2.5%) | 157 (3%) | 85 (1.8%) |  |  |
| White | 4002 (88.6%) | 4200 (81.6%) | 4165 (88.8%) |  |  |
| BMI category (n, %) |  |  |  |  |  |
| Normal or underweight | 37 (0.8%) | 923 (20.0%) | 385 (8.2%) | 51.5 pp range | <2e-16 |
| Obese | 3895 (86.3%) | 1789 (34.8%) | 2429 (51.8%) |  |  |
| Overweight | 487 (10.8%) | 2403 (46.7%) | 1818 (38.8%) |  |  |
| Sleep duration (n, %) |  |  |  |  |  |
| 7-8 hours | 2266 (50.2%) | 3062 (59.5%) | 2818 (60.1%) | 10.7 pp range | <2e-16 |
| <7 | 1777 (39.4%) | 1611 (31.3%) | 1345 (28.7%) |  |  |
| >=9 | 440 (9.7%) | 444 (8.6%) | 515 (11%) |  |  |
| Smoking status (n, %) |  |  |  |  |  |
| Current smoker | 620 (13.7%) | 549 (10.7%) | 405 (8.6%) | 8.5 pp range | <2e-16 |
| Never smoked | 1938 (42.9%) | 2488 (48.3%) | 1981 (42.2%) |  |  |
| Previous smoker | 1882 (41.7%) | 2039 (39.6%) | 2254 (48.1%) |  |  |
| Alcohol status (n, %) |  |  |  |  |  |
| Daily drinker | 919 (20.4%) | 1549 (30.1%) | 1500 (32%) | 11.6 pp range | <2e-16 |
| Moderate drinker | 968 (21.4%) | 1212 (23.5%) | 1057 (22.5%) |  |  |
| Non-drinker | 834 (18.5%) | 803 (15.6%) | 724 (15.4%) |  |  |
| Social drinker | 1750 (38.8%) | 1538 (29.9%) | 1388 (29.6%) |  |  |
| Lipid lowering medication (n, %) |  |  |  |  |  |
| No | 1016 (22.5%) | 1096 (21.3%) | 954 (20.3%) | 2.2 pp range | 0.0403 |
| YES | 3499 (77.5%) | 4052 (78.7%) | 3736 (79.7%) |  |  |
| Blood pressure lowering medication (n, %) |  |  |  |  |  |
| No | 1417 (31.4%) | 2067 (40.2%) | 1454 (31%) | 9.1 pp range | <2e-16 |
| Yes | 3098 (68.6%) | 3081 (59.8%) | 3236 (69%) |  |  |
| Oral glucose lowering medication (n, %) |  |  |  |  |  |
| No | 1388 (30.7%) | 1846 (35.9%) | 1758 (37.5%) | 6.7 pp range | 1.25E-11 |
| Yes | 3127 (69.3%) | 3302 (64.1%) | 2932 (62.5%) |  |  |
| Insulins (n, %) |  |  |  |  |  |
| No | 3588 (79.5%) | 4512 (87.6%) | 4071 (86.8%) | 8.2 pp range | <2e-16 |
| Yes | 927 (20.5%) | 636 (12.4%) | 619 (13.2%) |  |  |
| Diet category (n, %) |  |  |  |  |  |
| High | 1466 (32.5%) | 1830 (35.5%) | 1623 (34.6%) | 4.7 pp range | 2.58E-06 |
| Intermediate | 1675 (37.1%) | 1913 (37.2%) | 1861 (39.7%) |  |  |
| Low | 1374 (30.4%) | 1405 (27.3%) | 1206 (25.7%) |  |  |
| PRS for T2DM tertile (n, %) |  |  |  |  |  |
| 1 | 1405 (31.1%) | 1703 (33.1%) | 1526 (32.5%) | 2.4 pp range | 1.18E-09 |
| 2 | 1477 (32.7%) | 1630 (31.7%) | 1526 (32.5%) |  |  |
| 3 | 1426 (31.6%) | 1705 (33.1%) | 1502 (32%) |  |  |
| Missing | 207 (4.6%) | 110 (2.1%) | 136 (2.9%) |  |  |
| APOE4 Carrier (n, %) |  |  |  |  |  |
| Missing | 924 (21.3%) | 812 (16%) | 815 (17.8%) | 5.3 pp range | 2.66E-09 |
| No | 2525 (58.2%) | 3096 (61.1%) | 2766 (60.4%) |  |  |
| Yes | 889 (20.5%) | 1158 (22.9%) | 998 (21.8%) |  |  |

Values are presented as mean (SD) for continuous variables and n (%) for categorical variables. For skewed continuous variables (HbA1c, CRP, MET), values are shown as median (IQR). p-values are from Kruskal-Wallis tests for continuous variables and $\chi^2$ tests for categorical variables. Effect sizes are reported as the maximum absolute standardized mean difference (|d|max) across clusters for continuous variables and as the absolute percentage-point range across clusters for categorical variables. Statistical tests are descriptive only, as several variables (including HbA1c, BMI, insulin
resistance) contributed to the clustering process; therefore, differences observed in these variables are expected and should not be interpreted as independent associations. eGFR values are expressed in mL/min/1.73m<sup>2</sup>; descriptive statistics were verified to ensure correct unit scaling.
Abbreviations: SOID, severe obesity-related inflammatory type 2 diabetes mellitus; MMD, mild metabolic type 2 diabetes mellitus; MARD-H, mild age-related type 2 diabetes mellitus with hypertension; HbA1c, glycated hemoglobin; MET, Total Metabolic Equivalent Task minutes per week for all physical activity; DBP, diastolic blood pressure; SBP, systolic blood pressure; SD, standard deviation; IQR, interquartile range; SMD, standardized mean difference; HDL, high-density lipoprotein cholesterol; LDL, low-density lipoprotein cholesterol; TG, triglycerides; eGFR, estimated glomerular filtration rate; BMI, body mass index; T2DM, type 2 diabetes mellitus; PRS, polygenic risk score; APOE, apolipoprotein E.

As shown in *Figure 1*, three distinct and reproducible subtypes were identified: severe obesity-related inflammatory diabetes (SOID; n = 4,515, 31.5%), mild metabolic diabetes (MMD; n = 5,148, 35.9%), and mild age-related hypertension-predominant diabetes (MARD-H; n = 4,690, 32.7%). Participants in the SOID cluster had markedly higher BMI (mean 36.6 kg/m^2^), severe insulin resistance, elevated CRP, and higher HbA1c than participants in the other clusters. The MMD cluster showed the most favorable metabolic profile, with the lowest HbA1c and SBP, representing the clinically mild reference phenotype. The MARD-H cluster comprised the oldest participants (mean age at diagnosis 61.6 years) with intermediate glycemia but an elevated SBP profile. Baseline characteristics of the MRI sub-cohort are shown in *Supplementary Table S4*.

**Figure 1.**
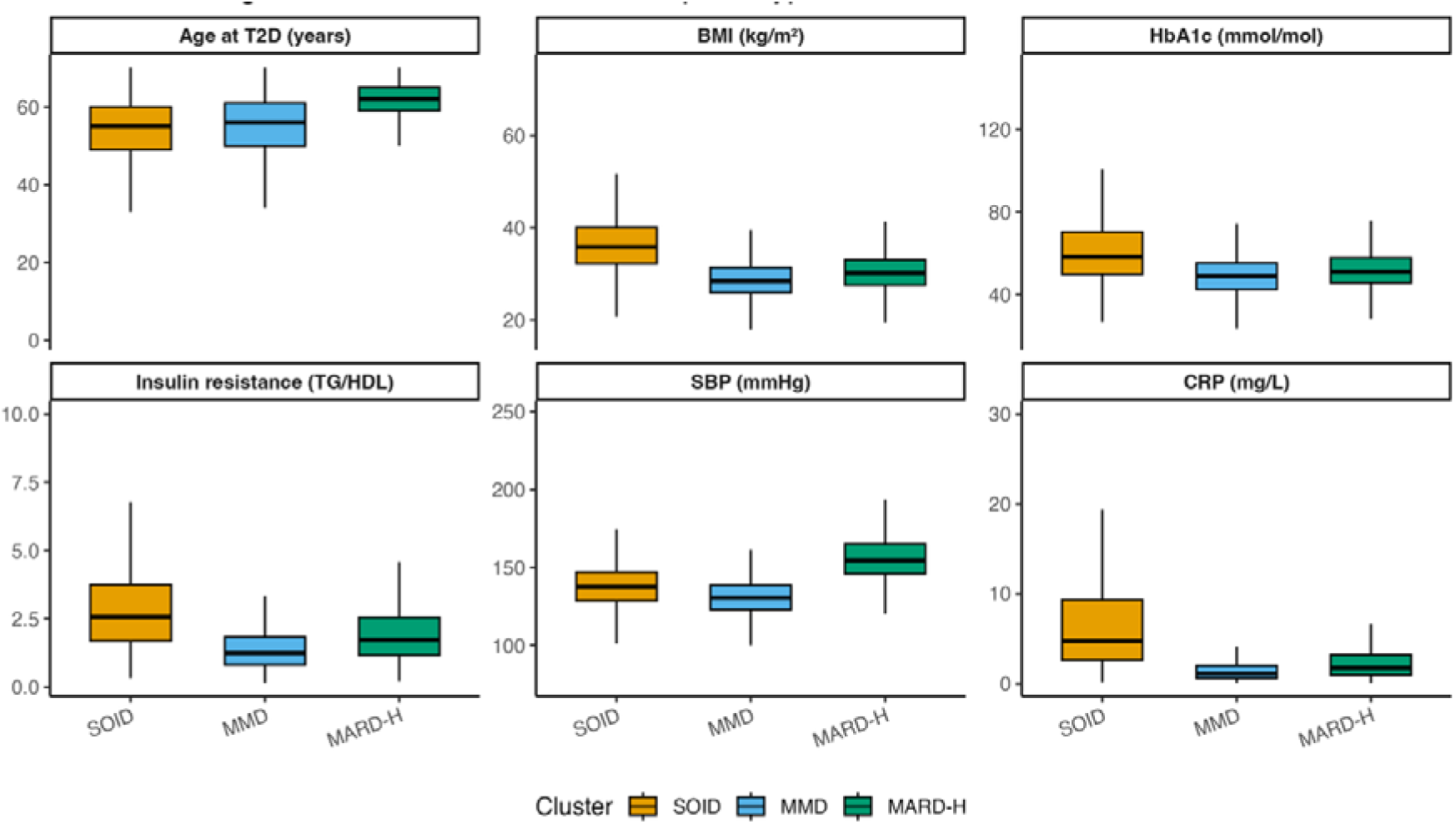
Distribution of cluster-defining variables across type 2 diabetes mellitus subtypes Boxplots show the median and interquartile range for each variable across the three T2DM subtypes. Insulin resistance was estimated using the triglyceride-to-HDL cholesterol ratio. All values shown are original (untransformed). Abbreviations: T2DM, type 2 diabetes mellitus; SOID, severe obesity-related inflammatory diabetes; MMD, mild metabolic diabetes; MARD-H, mild age-related hypertension-predominant diabetes; BMI, body mass index; HbA1c, glycated hemoglobin; TG, triglycerides; HDL, high-density lipoprotein; SBP, systolic blood pressure; CRP, C-reactive protein.

### 3.2 Associations with dementia outcomes

Associations between the T2DM clusters and dementia outcomes differed by subtype (*Figure 2*). For all-cause dementia, compared with the reference MMD cluster, the SOID cluster was associated with a higher hazard (HR = 1.24, 95% CI 1.01-1.52), whereas no increased hazard was observed for the MARD-H cluster (HR = 0.91, 95% CI 0.76-1.09). For AD, neither SOID (HR = 0.96, 95% CI 0.69-1.34) nor MARD-H (HR = 0.84, 95% CI 0.64-1.11) was statistically significantly associated with hazard after full confounder adjustment. In contrast, for VaD, the SOID cluster showed a significantly elevated hazard relative to MMD (HR = 1.42, 95% CI 1.01-1.99), while the association for MARD-H was weaker and not statistically significant (HR = 1.10, 95% CI 0.81-1.49).

**Figure 2.**
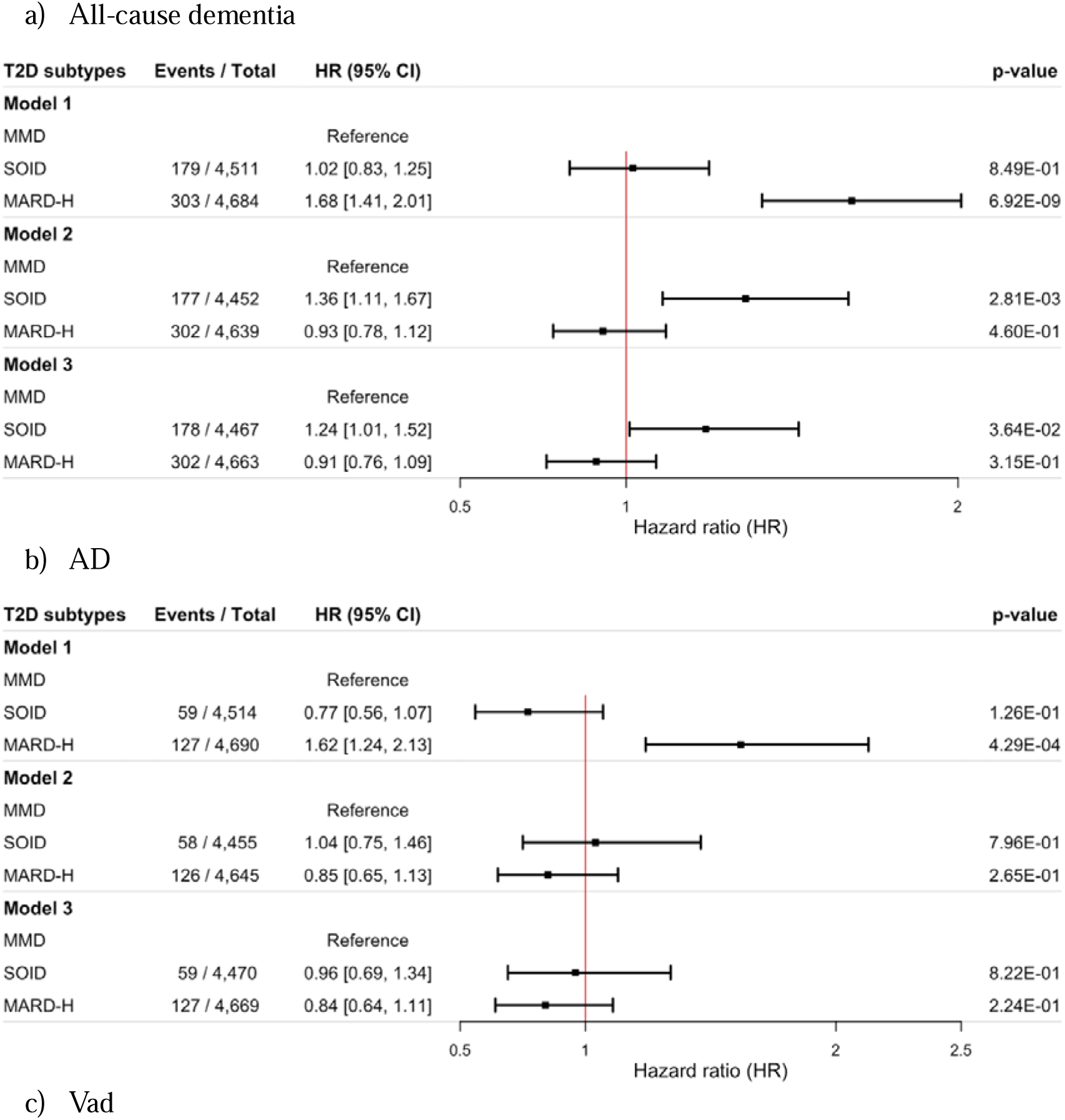
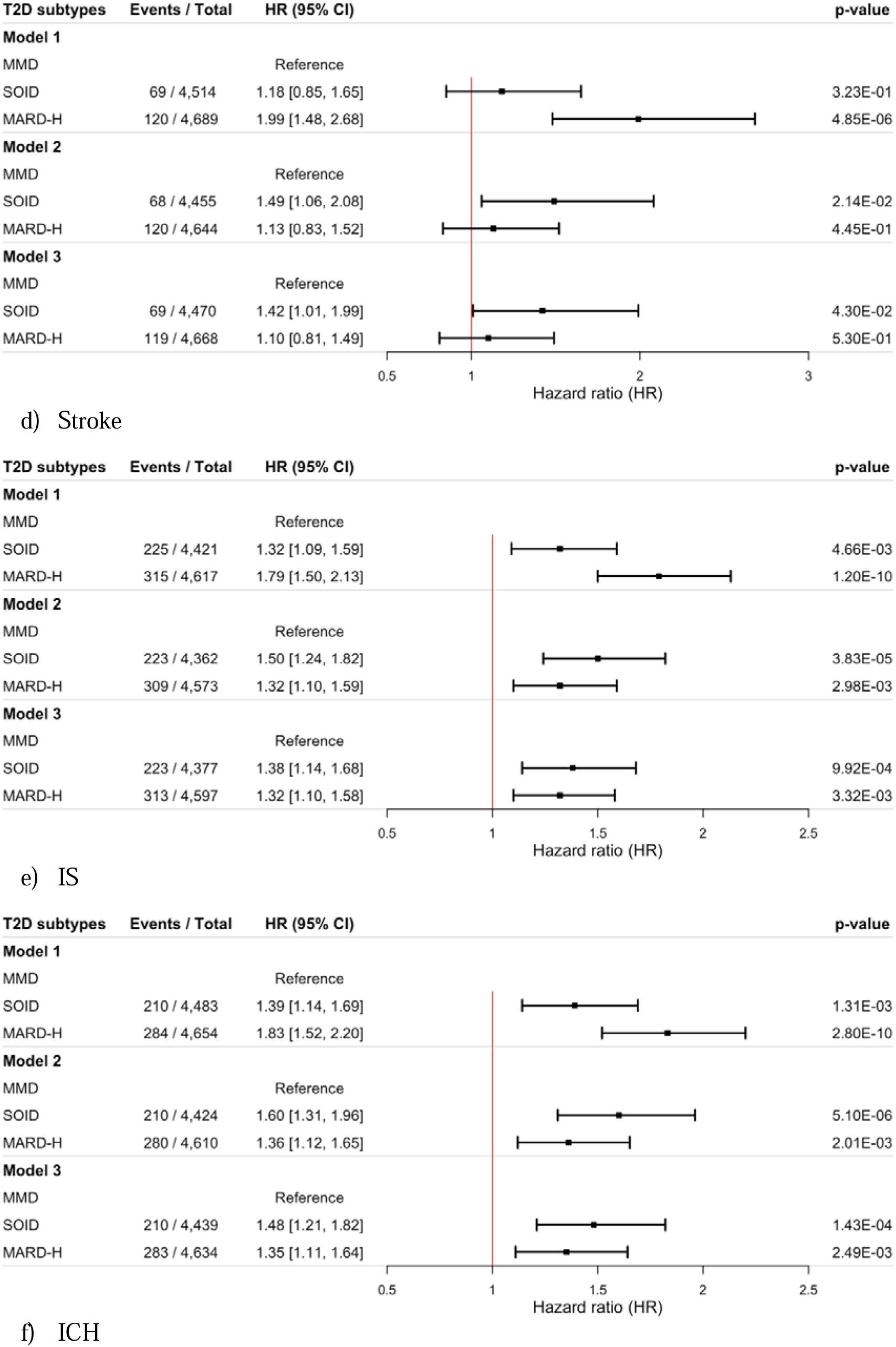
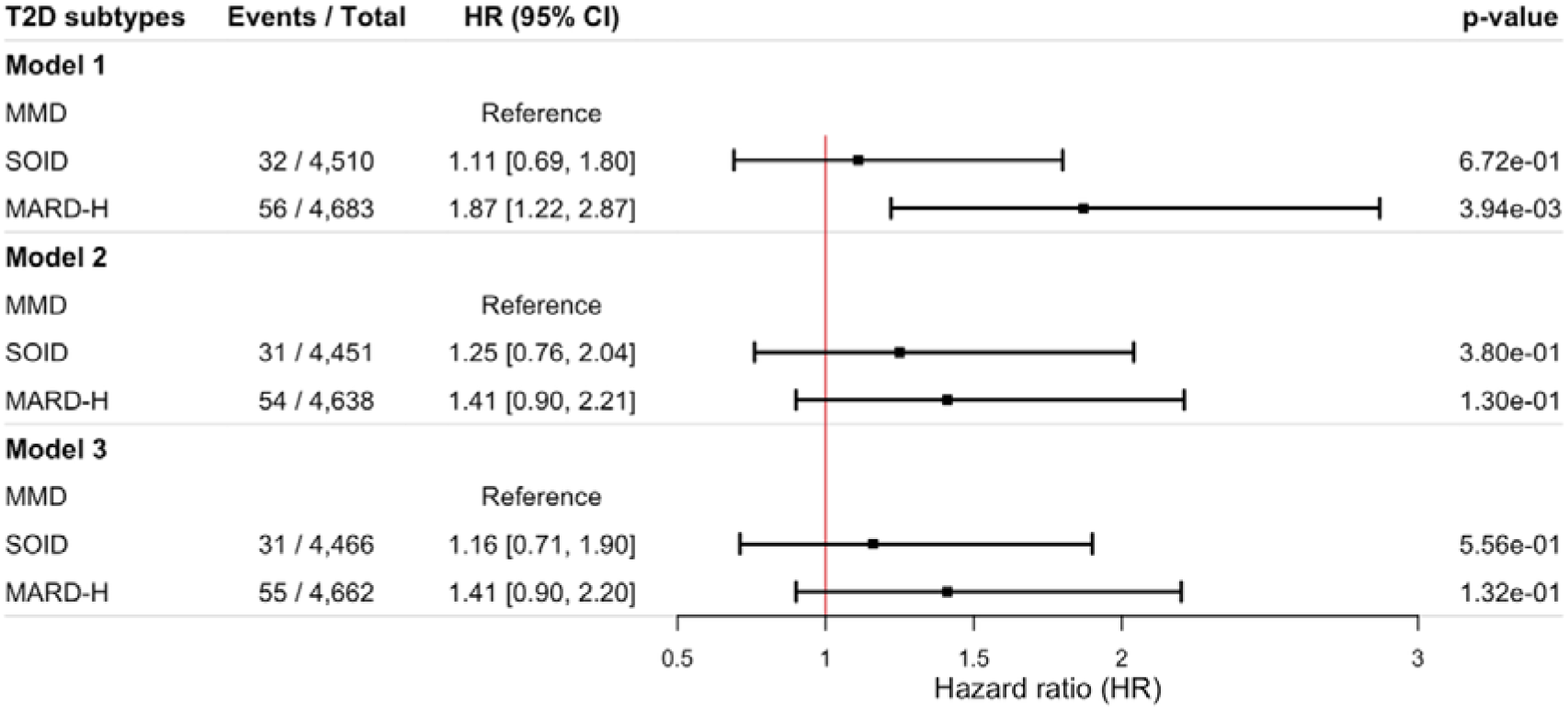
Associations between type 2 diabetes mellitus clusters and risk of dementia and stroke Hazard ratios (HRs) and 95% confidence intervals (CIs) were estimated using Cox proportional hazards models, with the mild metabolic diabetes (MMD) cluster as the reference group. Model 1 was unadjusted. Model 2 was adjusted for age at baseline, sex, and ethnicity. Model 3 was further adjusted for educational attainment, socioeconomic deprivation, smoking status, alcohol consumption, and overall diet score. Abbreviations: T2MD, type 2 diabetes mellitus; SOID, severe obesity-related inflammatory diabetes; MMD, mild metabolic diabetes; MARD-H, mild age-related hypertension-predominant diabetes; AD, Alzheimer’s disease; VaD, vascular dementia; IS, ischemic stroke; ICH, intracerebral hemorrhage; HR, hazard ratio; CI, confidence interval.

Although relative hazards were highest in the SOID cluster, cumulative incidence was highest in the MARD-H cluster, reflecting its older age distribution and higher baseline risk. For all-cause dementia, the 10-year cumulative incidence was highest in the MARD-H cluster (2.9%), compared with SOID (1.8%) and the reference MMD cluster (1.7%). RMST analyses were concordant, with participants in the MARD-H cluster experiencing fewer event-free years over 10 years (RMST 9.91 vs 9.95 years in MMD), while RMST was similar between SOID and MMD. For AD and VaD, absolute risks were lower overall but again tended to be highest in the MARD-H cluster, with small differences in RMST across clusters (approximately 9.97-9.99 event-free years over 10 years).

In analyses with additional adjustment for kidney function and medication use (Model 4), associations were partially attenuated, particularly for the MARD-H cluster, while estimates for SOID were less affected.

### 3.3 Associations with stroke outcomes

In confounder-adjusted Cox models (Model 3), risks of stroke outcomes varied across T2DM clusters (*Figure 2*). For all strokes, both non-reference clusters exhibited higher hazards compared with the reference MMD cluster, with the strongest association observed for SOID (HR = 1.38, 95% CI 1.14-1.68), followed by MARD-H (HR = 1.32, 95% CI 1.10-1.58). Similar patterns were observed for IS, where the SOID cluster showed a 48% higher risk (HR = 1.48, 95% CI 1.21-1.82) and the MARD-H cluster a 35% higher risk (HR = 1.35, 95% CI 1.11-1.64), relative to MMD. Neither the SOID cluster (HR = 1.16, 95% CI 0.71-1.90) nor the MARD-H cluster (HR = 1.41, 95% CI 0.90-2.20) showed a statistically significant association with ICH.

Over 10 years of follow-up, the cumulative incidence of stroke was highest in the MARD-H cluster (4.3%), followed by SOID (3.9%) and MMD (3.1%). RMST analyses were consistent with these patterns: compared with MMD, participants in the MARD-H cluster experienced a reduction of 0.07 event-free years over 10 years, while those in the SOID cluster experienced a reduction of 0.05 years. For IS, cumulative incidence was again highest in MARD-H (3.6%), followed by SOID (3.2%) and MMD (2.6%), with corresponding RMST losses of 0.06 and 0.05 event-free years for MARD-H and SOID, respectively. In contrast, ICH was rare across all clusters, with minimal differences in cumulative incidence and RMST.

With additional adjustment for kidney function and medication use, associations with stroke outcomes were attenuated. For any stroke, attenuation was modest for SOID (approximately 18%) and more pronounced for MARD-H (around 35%). Similar attenuation patterns were observed for IS and ICH.

### 3.4 Associations with mortality

Associations between T2DM clusters and risk of death varied by cause and level of covariate adjustment (*Figure 3*). In confounder-adjusted models (Model 3), the SOID cluster was consistently associated with a higher risk of all-cause mortality (HR = 1.59, 95% CI 1.45-1.74), cardiovascular mortality (HR = 1.88, 95% CI 1.59-2.23), and non-cardiovascular mortality (HR = 1.41, 95% CI 1.26-1.57), compared with the reference MMD cluster. In contrast, associations for the MARD-H cluster were substantially attenuated after confounder-adjustment for all-cause mortality (HR = 1.02, 95% CI 0.94-1.12), cardiovascular mortality (HR = 1.17, 95% CI 0.99-1.39), or non-cardiovascular mortality (HR 0.95, 95% CI 0.85-1.06).

**Figure 3.**
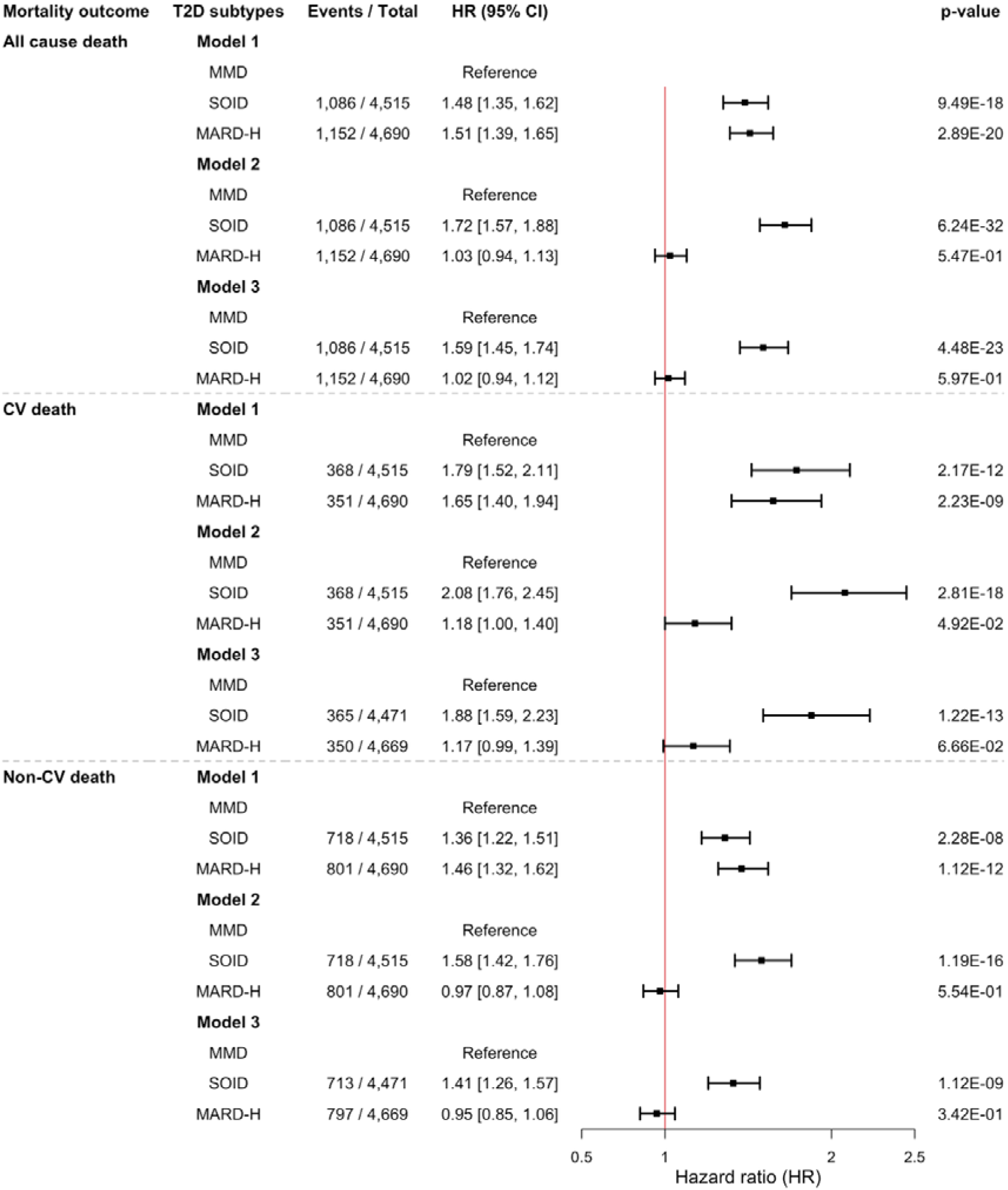
Associations of type 2 diabetes mellitus clusters with cause-specific mortality Hazard ratios (HRs) and 95% confidence intervals (CIs) were estimated using Cox proportional hazards models, with the mild metabolic diabetes (MMD) cluster as the reference group. Model 1 was unadjusted. Model 2 was adjusted for age at baseline, sex, and ethnicity. Model 3 was further adjusted for educational attainment, socioeconomic deprivation, smoking status, alcohol consumption, and overall diet score. Abbreviations: T2DM, type 2 diabetes mellitus; SOID, severe obesity-related inflammatory diabetes; MMD, mild metabolic diabetes; MARD-H, mild age-related hypertension-predominant diabetes; HR, hazard ratio; CI, confidence interval; CV, cardiovascular.

### 3.5 Brain structural differences

In confounder-adjusted (Model 3) IPW analyses, associations between T2DM clusters and brain MRI measures differed by subtype and imaging phenotype (*Figure 4*). Compared with the MMD cluster, the SOID cluster was associated with lower gray matter volume (standardized β = −0.20, 95% CI −0.34 to −0.06; p=0.021) and greater WMH burden (β = 0.33, 95% CI 0.16 to 0.49; p=9.16×10^-4^). No significant differences were observed for total brain volume, white matter volume, or hippocampal volume in the SOID cluster. In contrast, the MARD-H cluster showed a higher white matter volume compared with MMD (β = 0.23, 95% CI 0.05 to 0.41; p=0.047), while associations with total brain volume, gray matter volume, WMH burden, and hippocampal volume were not statistically significant.

**Figure 4.**
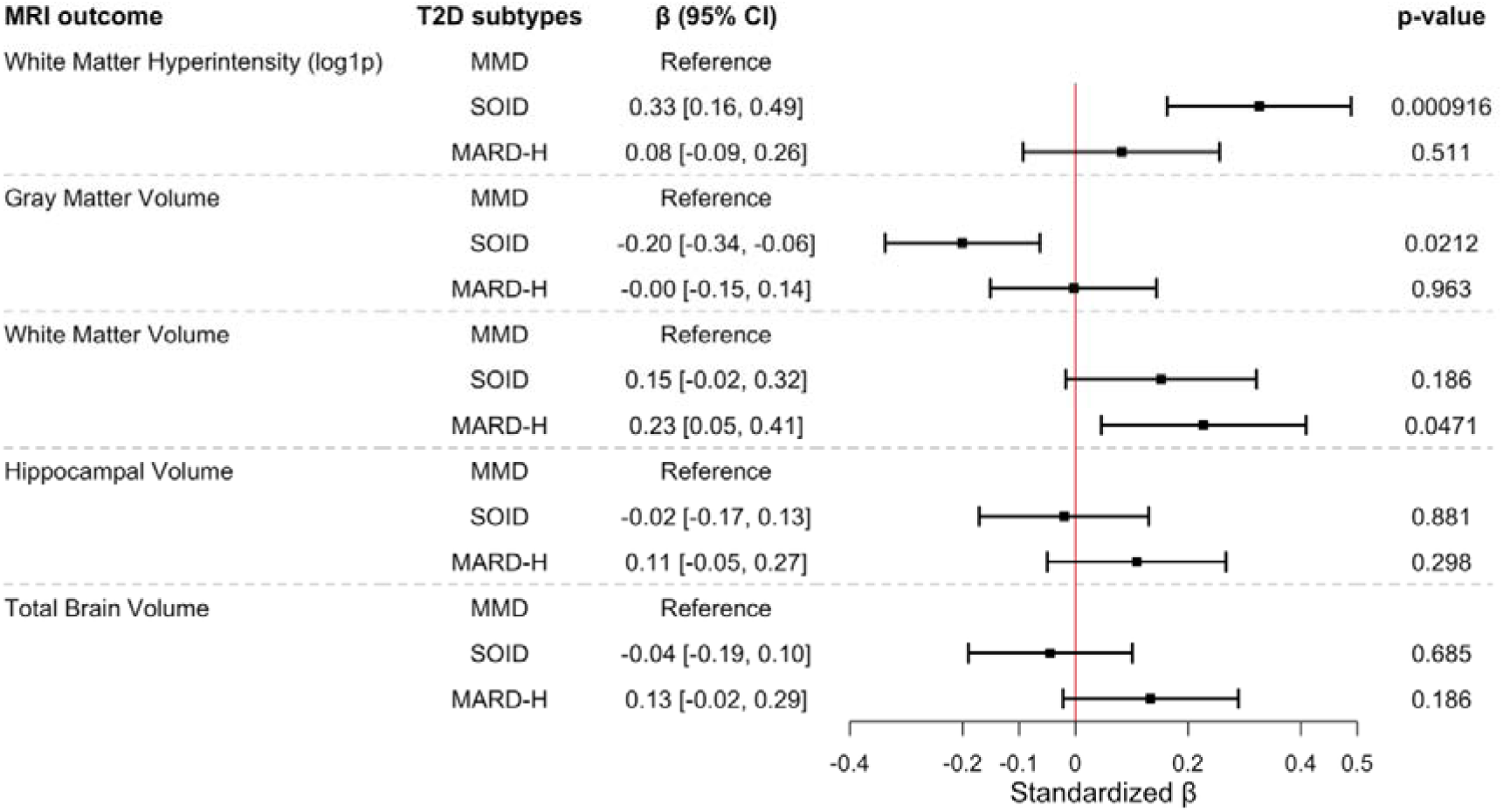
Associations between type 2 diabetes mellitus clusters and brain MRI measures (confounder-adjusted model) Standardized β coefficients and 95% confidence intervals were estimated using inverse probability–weighted linear regression models to account for non-random participation in brain MRI. Models were adjusted for confounders including age at baseline, sex, ethnicity, educational attainment, socioeconomic deprivation, smoking status, alcohol consumption, diet quality, and intracranial volume. The mild metabolic diabetes (MMD) cluster served as the reference group. Adjusted p-values are provided for descriptive purposes. Abbreviations: T2DM, type 2 diabetes mellitus; SOID, severe obesity-related inflammatory diabetes; MMD, mild metabolic diabetes; MARD-H, mild age-related hypertension-predominant diabetes; MRI, magnetic resonance imaging; β, standardized regression coefficient; CI, confidence interval.

### 3.6 Sensitivity analysis

Results were robust in multiple sensitivity analyses. In the complete-case dataset (n = 11,600; 80.8% of the analytic sample), the same three-cluster structure was identified, supported by consistent internal validation metrics (mean Jaccard 0.78-0.85). Cluster profiles closely matched those from the main imputed analysis, confirming reproducibility of the derived subtypes. Detailed characteristics and stability diagnostics are presented in *Supplementary Figures S6-S8*. After excluding the five-year diagnosis cases of dementia and stroke, the associations were consistent with the main results. In fully adjusted models (Model 3), SOID was associated with higher risks of all-cause dementia (HR = 1.27, 95% CI 1.01-1.59), VaD (HR = 1.57, 95% CI 1.08-2.06), stroke (HR = 1.46, 95% CI 1.18-1.82), and IS (HR = 1.55, 95% CI 1.24-1.94), while MARD-H showed elevated risks for several outcomes, including stroke (HR = 1.35, 95% CI 1.1-1.66) and IS (HR = 1.35, 95% CI 1.09-1.68). Estimates for ICH were imprecise due to the limited events.

When restricting the sample to participants with diabetes duration ≤1 year at baseline, associations were broadly consistent with the primary analyses, though with wider confidence intervals. For dementia outcomes, compared with the reference MMD cluster, the SOID cluster showed a higher risk of VaD (HR 2.14, 95% CI 1.12-4.10), whereas associations with all-cause dementia (HR 1.41, 95% CI 0.95-2.09) and AD (HR 0.89, 95% CI 0.48-1.65) were not statistically significant. Associations for the MARD-H cluster were generally attenuated and not statistically significant. For stroke outcomes, the SOID cluster remained associated with higher risks of stroke (HR 1.68, 95% CI 1.17-2.41) and IS (HR 1.76, 95% CI 1.20-2.58). Associations for ICH were elevated but imprecise (HR 2.77, 95% CI 0.99-7.75) (*Supplementary Table S6)*.

In Fine-Gray subdistribution hazard models accounting for death as a competing event, associations for stroke outcomes remained largely consistent with the primary analyses, whereas dementia associations were attenuated. Compared with the reference MMD cluster, the SOID cluster showed higher risks of stroke (sHR 1.32, 95% CI 1.09-1.60) and IS (sHR 1.40, 95% CI 1.14-1.71), and the MARD-H cluster showed similar elevations (stroke: sHR 1.34, 95% CI 1.11-1.61; IS: sHR 1.36, 95% CI 1.12-1.65). Associations for ICH were directionally similar but not statistically significant. In contrast, associations with all-cause dementia, AD, and VaD were attenuated and no longer statistically significant after accounting for competing mortality (*Supplementary Table S7)*.

## Discussion

In this large population-based cohort of 14,353 individuals with T2DM, we identified three reproducible clinically derived diabetes subtypes using routinely measured variables and demonstrated that these subtypes differ in their associations with dementia, stroke, mortality, and brain structural measures. While previous clustering structure broadly resembles diabetes phenotypes and has primarily focused on vascular complications (8, 14–16), this study extends subtype characterization to dementia subtypes, stroke subtypes, and structural neuroimaging markers within a unified framework. Our findings extend prior work by showing that these familiar clinical profiles are associated with *distinct neurological risk patterns*, including divergence between vascular and Alzheimer-type dementia and between relative and absolute risk estimates.

The clustering approach used in this study relied exclusively on routinely available clinical measures, facilitating comparability with prior work and potential translational relevance. The identified subtypes showed good internal reproducibility and reflect clinically meaningful dimensions of diabetes heterogeneity, including metabolic control (HbA1c, insulin resistance), adiposity (BMI), blood pressure burden (SBP), systemic inflammation (CRP), and disease onset (age at diagnosis). These dimensions have consistently been identified in prior data-driven T2DM subtyping studies as yielding reproducible diabetes subtypes with distinct cardiometabolic risk profiles, capturing the major biological axes of T2DM heterogeneity (7, 15, 17–19).

First, the subtype characterized by higher adiposity, a higher triglyceride-to–HDL cholesterol ratio, elevated CRP, and poorer glycemic control (SOID cluster) showed consistently higher relative risks of vascular outcomes, including VaD, IS, and cardiovascular mortality, compared with the reference MMD cluster. This pattern aligns with prior T2DM clustering studies reporting higher macrovascular complication rates and mortality among insulin-resistant or poorly controlled diabetes subtypes, supporting the notion that chronic hyperglycemia and insulin deficiency are key drivers of cerebrovascular and cardiovascular risk (8, 15). Importantly, this subtype was not associated with AD, suggesting that the excess dementia risk observed in this group is more strongly linked to vascular rather than neurodegenerative mechanisms. This distinction is clinically meaningful, as many previous studies examining diabetes-related dementia risk have not differentiated between dementia subtypes, potentially masking important heterogeneity in the underlying pathways (20, 21).

Our findings further extend this evidence by demonstrating corresponding alterations in brain structure, including greater white matter hyperintensity burden and lower gray matter volume, suggesting a potential neurological substrate underlying the elevated dementia risk observed in this cluster. In contrast, the older-onset subtype with higher systolic blood pressure exhibited modest relative hazards for dementia but the highest absolute risks of dementia and stroke over 10 years of follow-up.

This divergence between relative and absolute risk reflects the older age distribution and higher baseline risk in this group, underscoring the importance of reporting absolute risk metrics alongside hazard ratios, particularly in older populations with competing mortality. From a clinical perspective, these findings highlight that subtypes with modest relative risks may still account for a substantial burden of events, reinforcing the need to consider cumulative risk when assessing prognosis and prioritizing prevention strategies (8). For stroke outcomes, both non-reference subtypes were associated with higher risks of any stroke and ischemic stroke, with stronger relative associations observed for the SOID cluster. Associations with intracerebral hemorrhage were weak and imprecise, likely reflecting limited event numbers and potentially distinct etiological pathways. Together, these results indicate that clinically derived diabetes subtypes capture heterogeneity in cerebrovascular risk that is not apparent when T2DM is treated as a single homogeneous condition (22, 23).

Additional adjustment for kidney function and medication use attenuated several associations, particularly for the older-onset subtype, suggesting that downstream clinical factors related to disease severity and comorbidity burden partially account for the observed risk patterns. These analyses were conducted to assess robustness rather than to infer mediation and should not be interpreted as evidence for specific causal pathways.

Mortality analyses further highlighted heterogeneity across diabetes subtypes. The SOID cluster was associated with substantially higher risks of all-cause, cardiovascular, and non-cardiovascular mortality, consistent with previous observations that metabolically severe diabetes phenotypes experience poorer long-term survival (24). In contrast, excess mortality risk in the older-onset subtype was largely attenuated after adjustment, indicating that age and comorbidity burden, rather than subtype-specific effects, explain much of the observed mortality risk.

Competing risk analyses indicated that stroke associations were robust to accounting for death, with similar effect estimates in Fine-Gray and Cox models. In contrast, dementia associations particularly for the SOID cluster were attenuated after accounting for competing mortality, suggesting that excess death risk may partially obscure cumulative dementia risk. Given the substantially higher cardiovascular and all-cause mortality in SOID, differential survival likely influences observed dementia incidence. These findings underscore the importance of considering competing mortality when evaluating long-term neurological outcomes across diabetes phenotypes.

In exploratory brain MRI analyses, the SOID cluster showed greater white matter hyperintensity burden and lower grey matter volume compared with the metabolically mild subtype. In contrast, the older-onset subtype showed higher white matter volume. Although these findings are directionally consistent with the observed vascular risk profiles, the imaging analyses were cross-sectional and conducted in a selected subgroup of survivors, with a relatively limited sample size and potential residual selection bias despite inverse probability weighting. Accordingly, these results should be interpreted as descriptive and hypothesis-generating rather than indicative of temporal or causal relationships linking diabetes subtypes to brain structural changes.

Key strengths of this study include the large, well-characterized cohort, use of an unsupervised learning approach minimizing a priori bias, availability of brain imaging data, and integration of genetic information. The clustering variables were all routinely available in clinical practice, enhancing translational relevance. However, several limitations merit consideration. First, although we adjusted for a wide range of covariates, residual confounding cannot be excluded. Second, cluster assignments were based on baseline data and may not capture dynamic metabolic changes over time. Third, dementia diagnoses were ascertained from clinical and registry data, which may underestimate mild cognitive impairment or subclinical pathology. Fourth, external validation in diverse populations is needed to confirm generalizability. Finally, the UK Biobank cohort is healthier and less ethnically diverse than the general population, which may limit generalizability of absolute risk estimates.

In conclusion, this study demonstrates that clinically derived subtypes of T2DM are associated with distinct patterns of vascular and neurological outcomes, including divergence between vascular and Alzheimer-type dementia and between relative and absolute risk metrics. These findings highlight the heterogeneity of long-term neurological risk in T2DM and emphasize the value of subtype-informed and absolute-risk–based approaches when considering prognosis and prevention. Further work is needed to validate these findings and to determine whether such subtyping can meaningfully inform clinical risk stratification.

## Author Contributions

F.A. and G.J.B. conceived and designed the study. S.H. conducted the statistical analyses under the supervision of F.A., G.J.B., and M.S.. S.H. performed the analysis and interpretation of the data. S.H. drafted the manuscript, and F.A., G.J.B., M.S., and Y.Z. critically reviewed the article for important intellectual content. All authors provided final approval of the version to be published.

## Availability of data and materials

This research was conducted using the UK Biobank resource under application number 100993. The UK Biobank data are available to bona fide researchers upon application and approval by UK Biobank (www.ukbiobank.ac.uk). Restrictions apply to the availability of these data, which were used under license for the current study and are not publicly available.

## Data Availability

All data produced in the present study are available upon reasonable request to the authors

